# Health Disparity and Geospatial Analysis among Cancer Patients in the US/Mexico Border Region of Imperial Valley

**DOI:** 10.1101/2025.07.07.25331030

**Authors:** Kyle Fontelera, Alonzo Mendoza, Carla Francisco, Samantha Hall, Harrison Yang, Sandip Patel, Helen Palomino, Eunjeong Ko, Ming-Hsiang Tsou

## Abstract

**Purpose:** Disparities in access to treatment and quality of care among cancer patients is an important issue that influences disease outcomes in minority populations. Although past research regarding the association between travel and health outcomes of cancer patients exists, studies in rural regions such as the US-Mexico border are limited. The purpose of this study is to understand the spatial patterns and travel distance association on the health outcomes of various cancers in underserved rural regions in Southern California (Imperial Valley).

**Methods:** Patients were identified from the intake assessment data form for new patients in a community-based cancer organization (CBO) between 2006 and 2022. Analysis was conducted using ArcGIS Pro to visualize spatial patterns of cancers and to identify the relationship between travel distance and mortality. The relationship between ethnicity and cancer deaths were also investigated. Kernel Density Estimation (KDE) plots were used to visualize spatial patterns of certain groups.

**Results:** There is a significant association between ethnicity and cancer deaths. The odds of death are slightly higher among Hispanics/Latinos. As hospital distance increased, the odds of mortality decreased. There were no significant associations between travel distances and any of the stages of diagnosis and death. There were three types of spatial distribution patterns of cancers respective of the overall dataset.

**Conclusion:** Our study found three types of spatial distribution patterns of various cancers in Imperial Valley. Cancer patient mortality did not have statistical significance associations with travel distance to treatment. However, further investigation and better tools may yield clearer results regarding its correlation.

## Background

Addressing and eliminating healthcare disparities among minority populations is an increasingly prioritized area of research in public health.^1^ Disparities in cancer prevention, detection, treatment, and mortality rates stem from various factors including race/ethnicity, socio-economic status, and geographic location.^3–5^ For instance, Hispanics in the US are more likely than non-Hispanic whites to receive cancer diagnoses at later stages and exhibit higher incidences of specific cancer types such as liver, stomach, and cervical cancer.^6^ Cancer is the second leading cause of death among Hispanics with over 250 thousand deaths in 2022 and 2023.^2,9^ Cancer patients, often Hispanic, fall into the lowest quartile of income and educational attainment, further complicating their access to adequate care.^8^

Additionally, rural residents face disparities in cancer care, such as lower screening rates, later cancer detection, and poorer survival outcomes compared to urban counterparts.^11–13^ For example, rural areas show a higher distance to mammography centers and greater rates of breast cancer (BC) diagnoses at late stage compared to urban areas.^11^ Limited access to medical professionals, longer travel times, and sparse healthcare resources exacerbate these disparities.^11,14^ Rural cancer patients experience limited access to medical and oncology providers, longer travel times, and low recruitment to clinical trials.^14^ Rural areas experience higher mortality rates from cancer compared to urban areas, despite lower incidence rates.^6^ Rural-nonrural disparities occur throughout the United States, but are greatest in regions such as the South, Appalachia, tribal communities, and the US-Mexico border.^15^ For example, cancer patients who live along the Texas-Mexico border, where access to healthcare is suboptimal, have lower 5-year-survival rates.^7^

Rural America faces the most hospital closures, physician shortages, and an aging workforce. The percentage of oncologists practicing in rural areas remains around 15%. The geographic density of oncologists in rural areas is another concern. More than half of all counties in the United States have no available oncologists. In comparison, urban areas have 20 times the number of oncologists by land area.^15^ Approximately 20% of rural America lives greater than 60 miles from a medical oncologist. Travel distance is associated with negative clinical outcomes, such as later stage of diagnosis, less timely receipt of chemotherapy, and delaying or declining treatment.^15^ Increasing travel requirements are associated with a more advanced diagnosis, inappropriate treatment, worse prognosis, and decreased quality of life.^16^ Patients residing in rural regions or areas with poor geographic access were twice as likely to not know their cancer stage than their urban counterparts, suggesting that these patients do not receive full diagnostic evaluation. In a study of patients with rectal cancer, death was more likely in patients living farther from radiotherapy services, with a 6% increase in mortality risk found for each 50-mile increase in distance from the nearest facility.^16^

Previous studies have thoroughly documented travel burdens, but combined with a lack of financial resources, transportation and language barriers, complications arise and conflict with cancer care.^4,6,8,10^ Despite these factors, which are supported in scholarly literature, little is known about the extent of travel burdens that may cause delays in cancer diagnosis and receiving recommended cancer treatments. The aims of this study are to determine 1) whether the travel distance to treatment facilities is negatively associated with survival and 2) whether cancer disparities exist among Hispanic versus non-Hispanic whites living in Imperial Valley in Southern California, adjacent to the US-Mexico border, and 3) identify similarities and differences in spatial patterns among different cancer types and demographic characteristics.

## Methods

### Ethical statement

This study was approved by the University Institutional Review Board [Blinded for Peer Review]. Participant’s confidentiality is protected by ensuring removal of any identifiable patient information.

### Data Source

This is an open prospective cohort study and patients are followed closely until death or the end of the study. The data for this study was collected as part of the intake assessment for new cancer patients who registered for services at a non-profit cancer organization in the California, US-Mexico border region. Intake data were obtained during the first contact with patients when they initiated services related to their treatment plan. From 2006 to 2022, a total of 3116 patients’ intake information were electronically entered into the community-based cancer organization database. This dataset includes cancer patients’ sociodemographic (i.e., gender, age, marital status, race/ethnicity) and cancer related information (i.e., types and stage of cancer, place of cancer diagnosis, new or recurrent diagnosis, location of cancer clinics).

### Geospatial analysis

There were 3,116 observations in our original dataset, but some of our data were eliminated due to insufficient information regarding age, participant address, treatment center name, and geocoding issues, resulting in 2,365 remaining observations. Using a geocoding method and network analysis in ArcGIS Pro, we calculated travel distance utilizing the direct distance (a straight-line distance) from the patients’ homes to their respective treatment facilities and examined the effects of travel distance to mortality. We conducted a descriptive analysis by using Tableau (a data visualization software) and ArcGIS Pro (a GIS software).

### Health Disparity Statistical Analysis

A chi-square analysis was performed to assess the association between race/ethnicity and cancer-related mortality. The analysis focused on two main ethnic groups: Hispanic and Non-Hispanic White. We compared the distribution of deaths between Hispanic and Non-Hispanic Whites. The chi-square test assessed whether the observed distribution of deaths differed significantly from the expected distribution under the null hypothesis of no association. Additionally, the contingency table obtained from the analysis allowed us to calculate the odds ratio for Hispanic individuals’ cancer-related mortality relative to Caucasian individuals. Statistical analyses were performed using R software, and results were considered significant at a p-value of <0.05. We employed logistic regression, utilizing stepwise subset selection based on AIC, to investigate the factors influencing death outcomes (yes or no) among cancer patients. The primary predictor in our analysis was distance to cancer treatment center, which reflects individuals’ proximity to healthcare facilities. Covariates include ethnicity, socioeconomic status (determined by a combination of their income and how many individuals reside in their household), age, and cancer type, to comprehensively examine their impact. The logistic regression model enabled us to compute odds ratios and evaluate their statistical significance, providing insights into the associations between these predictors and the likelihood of death outcomes. The dataset was then stratified into four major cancer types: breast, lung, colorectal, and prostate. To investigate the factors associated with mortality within each cancer type, separate logistic regression analyses were conducted. For each subset, the primary outcome variable was the binary indicator of patient death (yes or no). The predictor variables considered were similar to the original model and included distance to healthcare facilities, patient age, and cancer stage. Logistic regression models were used to estimate the odds ratios (ORs) and their significance, highlighting the associations between these variables, and the likelihood of death for each cancer type. All statistical analyses were conducted using the R software environment, and results were deemed significant at a threshold p-value of <0.05.

## Results

### Demographics

The participants’ socio-demographic and health-related variables were described in Table 1. There were 2365 cancer patients in total. 54-74-year-olds made up almost half of the cancer patients of the study population, followed by 65-74-year-olds made up 30% of the study population. About 60% were female and the average age was 65 years. Most patients were Latino/Hispanic (84.06%). Breast cancer was the most common cancer diagnosis (29.7%), followed by Colorectal cancer (9.09%). A plurality of patients (n= 469, 26.32% of all study participants) were in their fourth stage of cancer upon enrollment. Among 15-39-year-olds, most are in stage 4 cancer while among 40-64-year-olds, most are in stage 2, followed closely by stage 3. As for individuals 75-years-olds and older, most are in stage 4, followed by stage 2. The most frequented hospitals visited were El Centro Regional Medical Center (ECRMC) Hematology and Oncology Center (n=573), Sharp in San Diego (n=483), and Esperanza Oncology (n=185). The average distance traveled to each hospital is 10.7km, 140.4km, and 15.4 km. ECRMC and Esperanza Oncology are local cancer treatment centers in Imperial Valley; however, Sharp is one of the most attended hospitals in this dataset and is considerably far compared to the other hospitals. Furthermore, the total distance accrued among all patients driving to Sharp is 67,700km for one visit. More than one thousand patients in this dataset use either ECRMC Oncology Clinic or Sharp as their primary hospital. USC Medical is the farthest hospital, with an average patient home-to-hospital distance of 290.4km. The next highest total distance is UCSD Thornton, at 13,000 kilometers. About 64% (n=1419) of Imperial Valley Cancer residents in this dataset receive Cancer care from a local hospital in Imperial Valley. San Diego County has the most patients outside of Imperial Valley, with 29% (n=662) of residents going there for treatment.

**Table 1.** Patients’ socio-demographic characteristics. Distance, Age, Race/Ethnicity, Cancer Diagnosis, Cancer Stage at Diagnosis, and Place of treatment stratified by sex.

| **Characteristics**<br>n= 2365 | **Female**<br>**n=1421 (60%)** | **Male**<br>**n=944 (40%)** | **%** |
| --- | --- | --- | --- |
| **Distance(km) Median (Range)** | 17.6 (3 to 106) | 17.9 (1 to 102) | - |
| **Age- Average = 65.4** | Average= 64.4 | Average= 66.7 | - |
| 0-14 | 4 (33%) | 8 (67%) | .51% |
| 15-39 | 58 (50%) | 58 (50%) | 4.90% |
| 40-54 | 229 (73%) | 86 (27%) | 13.32% |
| 54-64 | 338 (71%) | 138 (29%) | 20.13% |
| 65-74 | 424 (60%) | 286 (40%) | 30.02% |
| 75-84 | 236 (52%) | 215 (48%) | 19.07% |
| 85+ | 132 (46%) | 153 (54%) | 12.05% |
| **Race/Ethnicity (n=2316)** |  |  |  |
| Latino/Hispanic | 1202 (60%) | 786 (40%) | 84.06% |
| Caucasian | 142 (55%) | 117 (45%) | 10.95% |
| African American | 20 (51%) | 19 (49%) | 1.65% |
| Asian/ Asia-Pacific | 12 (70%) | 5 (30%) | 0.72% |
| Other | 45 (73%) | 17 (27%) | 2.62% |
| **Cancer Diagnosis (n=2320)** |  |  |  |
| Breast | 687 (99.7%) | 2 (0.3%) | 29.70% |
| Colorectal | 92 (44%) | 119 (56%) | 9.09% |
| Prostate | 4 (2.1%) | 188 (98%) | 8.28% |
| Lung | 70 (46%) | 82 (54%) | 6.55% |
| Non-Hodgkin<br>Lymphoma | 55 (52%) | 51 (48%) | 4.57% |
| Leukemia | 26 (36%) | 47(64%) | 3.14% |
| **Cancer stage at diagnosis (n=1646)** |  |  |  |
| Stage 0 | 27 (90%) | 3 (10%) | 1.68% |
| Stage 1 | 167 (81%) | 40 (19%) | 11.62% |
| Stage 2 | 278 (76%) | 88 (24%) | 20.54% |
| Stage 3 | 178 (64%) | 99 (36%) | 15.54% |
| Stage 4 | 218 (53%) | 251 (54%) | 26.32% |
| Unknown | 325 | 258 |  |
| **Place of Treatment (n= 2229)** |  |  |  |
| Imperial Valley | 842 (59%) | 577 (41%) | 63.66% |
| San Diego | 402 (61%) | 260 (39%) | 29.70% |
| Riverside | 84 (63%) | 48 (36%) | 5.92% |
| Los Angeles | 8 (50%) | 8 (50%) | 0.72% |
| Missing | 85 | 2.16 |  |

In figure 1 we illustrate the average distance traveled to get to a health clinic, not just necessarily hospitals. We see travel from the Imperial Valley to as far as Los Angeles and San Diego. Additionally, shorter in distance but still relatively far from patients’ locations, Cathedral City and Indio are also popular locations to seek care.

**Figure 1.**
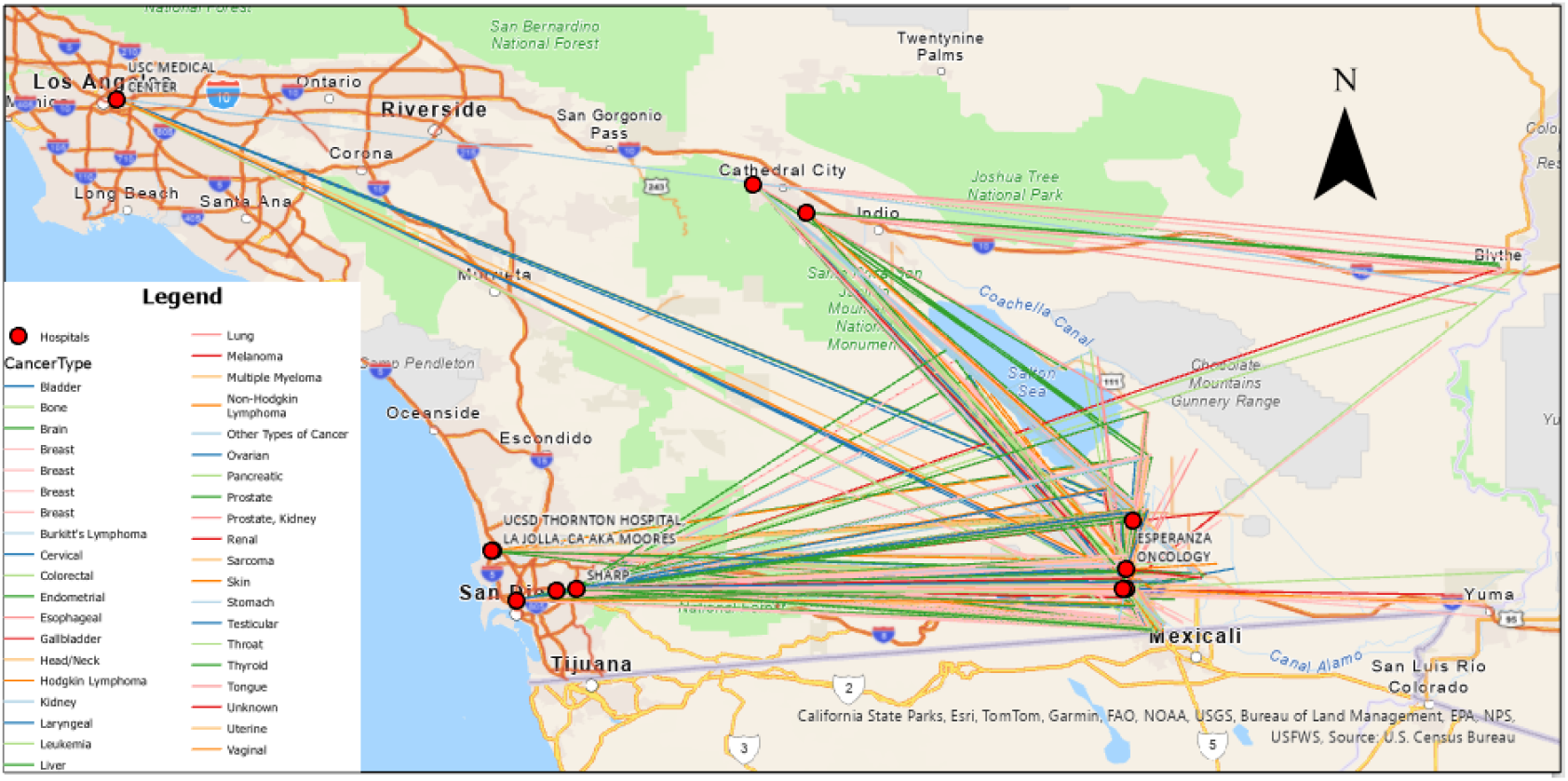
Map of straight-line distances from patients’ homes to their cancer treatment centers. Line colors represent the patients’ cancer types.

**Figure 2.**
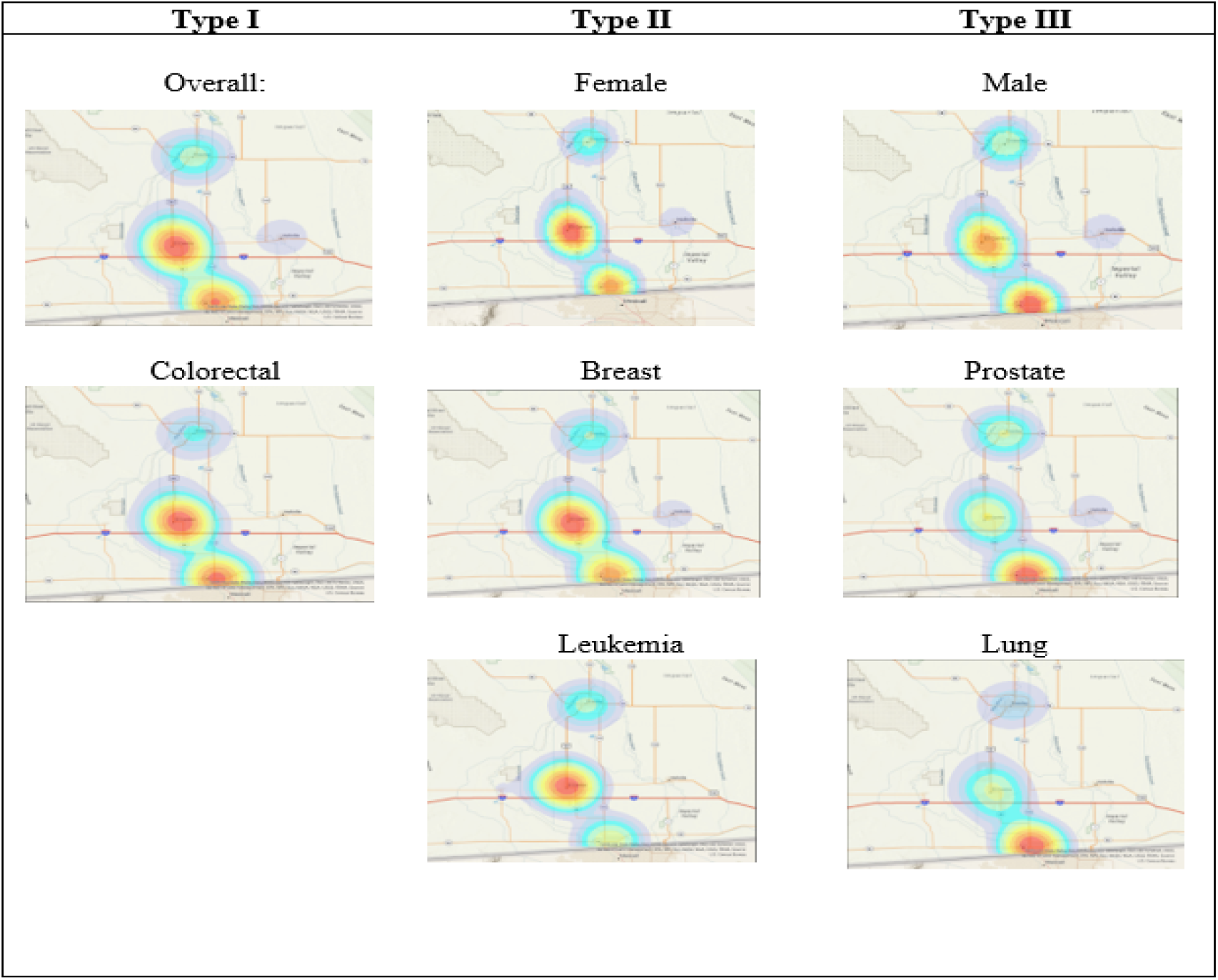
Spatial Patterns in Cancer patient concentration across the US/Mexico Border Region of Imperial Valley

### Comparison of study population to state and national

Table 2 presents the prevalence of cancer types in this study, comparing the state data in California and the United States data.^17^ To preserve continuity, the prevalence of cancer types is not written to avoid prevalence being confused with incidence rates in this data. The most prevalent cancer among women is Breast Cancer. Among non-Hispanic White (NHW) women, the second and third highest cancer counts in the study population are consistent with the state and national levels. Among men, the top 3 highest counts of cancer types (Prostate, Colorectal, & Lung) are consistent across the three levels and respective of whether they are NHW or Hispanic. The 4^th^ and 5^th^ positions among both Hispanic and non-Hispanic White women in this registry were not like the national nor the state.

**Table 2.** Comparison of the top 5 most prevalent cancers among males and females stratified by ethnicity. Ranking of the CBO dataset based on prevalence. Ranking of State and National based on incidence rates

|  | **CBO**<br>**patients in Imperial Valley** |  | **US (2020)<sup>17</sup>**<br>**Rates per 100,000** |  | **California (2020)<sup>17</sup>**<br>**Rates per 100,000** |  |
| --- | --- | --- | --- | --- | --- | --- |
| **Cancer Type**<br>**Rank**<br>**(Female)** | **Hispanic**<br>**(n= 1084)** | **NHW**<br>**(n= 134)** | **Hispanic** | **NHW** | **Hispanic** | **NH Whites** |
| 1. | Breast | Breast | Breast ( 86.2) | Breast (125.3) | Breast (87.0) | Breast (126.4) |
| 2. | Colorectal | Lung | Colorectal (24.1) | Lung (47.6) | Uterus (25.2) | Lung (35.2) |
| 3. | Lung | Colorectal | Uterus (23.4) | Colorectal (29.1) | Colorectal (24.4) | Colorectal (28.3) |
| 4. | NH Lymphoma | Ovarian | Lung (20.5) | Uterus (25.6) | Thyroid (16.6) | Melanoma (25.5) |
| 5. | Cervical | Melanoma | Thyroid (17.3) | Melanoma (21.9) | Lung and Bronchus (16.4) | Uterus (25.3) |
| **Cancer Type**<br>**Rank (Male)** | **Hispanic**<br>**(n= 786)** | **NHW**<br>**(n=117)** | **Hispanic** | **NHW (2020)** | **Hispanic** | **NHW** |
| 1. | Prostate | Prostate | Prostate (70.4) | Prostate (95.4) | Prostate (73.2) | Prostate (91.7) |
| 2. | Colorectal | Lung | Colorectal (32.8) | Lung (56.3) | Colorectal (33.0) | Lung (40.9) |
| 3. | Lung | Colorectal | Lung (28.0) | Colorectal (37.2) | Lung (21.9) | Melanoma (40.2) |
| 4. | NH Lymphoma | NH Lymphoma | Kidney & Renal Pelvis (20.6) | Urinary Bladder (33.5) | Kidney (20.9) | Colorectal (33.9) |
| 5. | Leukemia | Leukemia and Head/Neck | NH Lymphoma (17.5) | Melanoma (31.7) | NH Lymphoma (17.5) | Bladder (32.7) |

**Table 3.**
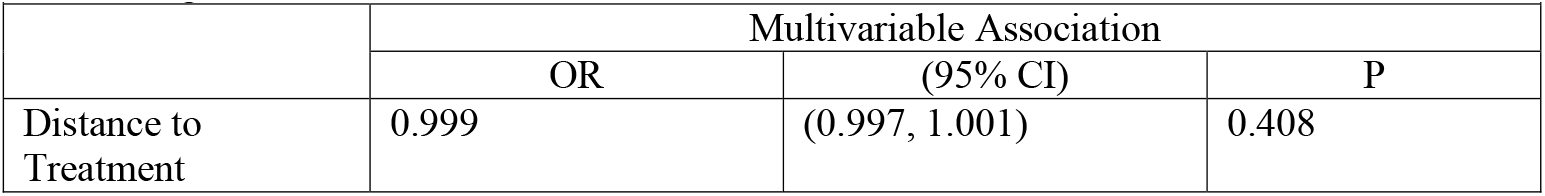
Multivariable Analysis of Distance to treatment and mortality, adjusting for age and cancer stage.

### Statistical Analysis Results

Among non-Hispanic whites, 69 individuals passed away, while 190 individuals survived from all cancers. Similarly, among Hispanics, 432 individuals died, and 1556 individuals survived from all cancers. The obtained chi-squared value was 2.9 with 1 degree of freedom, yielding a p-value of 0.08. With a significance level of 0.05, this suggests that there was no significant association between ethnicity and deaths from cancer. However, the odds of death from cancer for Hispanics were approximately 1.3 times higher relative to non-Hispanic whites. This suggests that Hispanics have a slightly elevated risk of cancer-related mortality in comparison to non-Hispanic Whites. Breast and Colorectal Cancer (for their high sample size), Prostate and Lung Cancer (for their unusual geographic patterns), after stratifying by Cancer Stage. For all strata, none yielded any significant results (p-value < 0.05). Investigating the bivariate associations between certain risk factors and mortality, we found that age and stage of cancer both had significant associations.

## Table

### GIS Mapping Analysis

The central kernel density represents the overall distribution encompassing all geocoded addresses within the database. Notably, the overall kernel density exhibits a prominent concentration of cancer patients in El Centro, with a smaller yet still significant density observed in Calexico. Three discernible patterns emerged, categorized as follows:

Type I: This pattern closely resembles the overall spatial distribution, characterized by two distinct high-density areas, notably featuring a central red spot. Colorectal Cancer displays a spatial distribution mirroring this overall kernel density plot.

Type II: This pattern exhibits similarities to the overall distribution, featuring a central circle of higher density juxtaposed with a less dense southern circle, devoid of intense red hues, often appearing orange or green. Notably, the high-density area within this pattern aligns with the top circle, situated at El Centro. Cancers such as Leukemia and Stomach Cancer demonstrate KDEs indicative of a notable concentration of cancer patients in El Centro, with a notable absence of high density in Calexico. These KDEs correspond to distributions representative of female-only cancer patients.

Type III: In stark contrast to the overall distribution and Type II, this pattern is characterized by a higher density of cancer patients in Calexico, with no significant density observed in El Centro. Prostate and Lung Cancer KDEs adhere to this pattern, aligning with distributions typical of male-only cancer patients.

In the figure above, we see that in the Type I spatial pattern, Colorectal Cancer’s spatial distribution closely resembles the overall kernel density plot. Among Females, in the Type II spatial pattern, Breast Cancer more closely follows the kernel density plot specifically with matching hot spot concentrations. The spatial distribution of Leukemia among females resembles it as well but with differences in density in the southern region. Finally, among males, prostate and lung cancer mostly resemble the Male cancer spatial distribution. The male prostate and lung cancer best mirror this spatial pattern especially in the southern region where concentrations are the highest.

## DISCUSSION

### Interpretation of Results

This study examines the association between the distance to treatment to care and mortality. Upon statistical adjustment for age and cancer treatment, our analysis revealed a marginal decrease of 0.01% in the odds of cancer-related mortality for each additional year of age. There was no significant association between the distance to treatment facilities and mortality rates. Nevertheless, subsequent post-hoc analysis revealed comparable differences in death proportions across hospitals in different counties, averaging around 22% (except in Los Angeles, where it reached 37.5%, potentially influenced by a small sample size). Furthermore, an unadjusted chi-square analysis failed to establish a significant relationship between mortality rates and hospital location. The distance analysis results were similar to a study done among Cervical Cancer patients where despite distance, recurrence rates of cancer were similar.^18^ Although our bivariate analysis predicted that the association is insignificant, this result is still peculiar because it contradicted similar studies which concluded that distance to care does exacerbate an individual’s condition, even leading to death.^16,19^ This may be because our sample sizes and demographics are different since this data set includes only those who sought assistance through the CBO. However, some studies have concluded that there is no association and others even found lower mortality for patients travelling longer distances for treatment.^20–22^ Regarding our study, there was no significant association between ethnicity and deaths from cancer. However, the odds of death from cancer for Hispanics were approximately 1.3 times higher relative to non-Hispanic whites. This suggests that Hispanics have a slightly elevated risk of cancer-related mortality in comparison to non-Hispanic Whites. Breast and Colorectal Cancer (for their high sample size), Prostate and Lung Cancer (for their unusual geographic patterns), after stratifying by Cancer Stage. For all strata, none yielded any significant results (p-value < 0.05).

### Geospatial Analysis

Following the generation of a series of Kernel Density plots, discernible spatial patterns emerged, categorized into three distinct types: overall (type I), female-specific (type II), and male-specific (type III). It was anticipated that the spatial distribution of cancer types would mirror the prevalence among the respective sexes. This hypothesis was corroborated for numerous cancers, including breast cancer, primarily affecting women, which exhibited spatial patterns akin to the female-specific distribution. Similarly, prostate cancer, prevalent among men, demonstrated spatial patterns aligning with the male-specific distribution. However, notable exceptions were observed, particularly in the case of lung cancer, which exhibits comparable incidence rates among males and females but displayed a distinct Type 3 spatial pattern contrary to expectations. Moreover, analysis revealed concentrated areas of high density, with El Centro exhibiting elevated female density and Calexico demonstrating a pronounced male concentration. Despite this, census data indicates a relatively balanced gender ratio in both locales, suggesting additional factors may underlie these observed patterns. Although stratification by gender allowed for the examination of spatial trends, the influence of these effects may vary across different geographical locations. Potential disparities in environmental factors promoting cancer incidence between the two cities warrant further investigation. Hence, these spatial patterns present a compelling avenue for future research to elucidate the interplay of environmental and geographical factors in cancer morbidity among Hispanics.

## Conclusion

The aims of this study are threefold: first, to examine whether the distance patients must travel to access treatment facilities is associated with mortality, secondly to assess potential disparities between Hispanic and non-Hispanic White populations in Imperial Valley, and finally to identify potential spatial patterns that may have links to different types of cancers and demographic characteristics. By investigating these factors, we aim to gain insights into the impact of travel distance on cancer care outcomes and to shed light on any inequities that may exist in accessing healthcare services among different demographic groups. We found no association between distance to treatment and mortality. However, there may be evidence that cancer types follow a spatial pattern based on specific strata. Through a kernel density estimation, we found that cancer types either followed an overall pattern (high concentration in both Calexico and El Centro), or specific sex (high concentration in El Centro for females and Calexico for males).

Strengths of this study include its utilization of a sizable dataset, encompassing a substantial portion of cancer patients residing in Imperial Valley. Moreover, this study represents the inaugural utilization of the CBO’s dataset, thus contributing to the advancement of research in this domain. Additionally, the investigation incorporates a spatial methodology, warranting further exploration. Nonetheless, limitations such as inaccurate records of addresses, dates, ages, and sample size changes must be acknowledged. Furthermore, possible outcome misclassification arises through methodology employed for identifying deaths, which included sources such as obituaries, familial communications, and cross-referencing with death registries. Consequently, there exists a probability that individuals categorized as alive within the dataset may be deceased. Moreover, this study is susceptible to selection bias, since it predominantly encompasses individuals who actively sought assistance from the CBO. Consequently, some of the cancer patient population not interfacing with the CBO remains unrepresented, thereby compromising the study’s overall representativeness. Finally, we also chose to use a simple regression method instead of also utilizing spatial statistical methods.

## Data Availability

All data produced in the present study are available upon reasonable request to the authors.

